# The Epidemic Severity Index: Estimating Relative Local Severity of Novel Disease Outbreaks

**DOI:** 10.1101/2020.04.23.20077685

**Authors:** Gareth Davies

## Abstract

Determining the severity of a novel pathogen in the early stages is difficult in the absence of reliable data. The pattern of outbreaks for COVID-19 across the globe have differed markedly above and below 30°N latitudes, suggesting very different levels of severity, but countries worldwide have implemented the same lockdown strategies. Existing methods for estimating severity appear not to have been useful in informing strategic decisions, possibly due to mismatches between the data required and those available, overly sophisticated methods with undesirable biases, or perhaps confusion and uncertainly generated by the wide range of estimates these methods produced early on.

The Epidemic Severity Index (ESI) is a simple, robust method for estimating the local severity of novel epidemic outbreaks using early and widely-available data and that does not depend on any estimated values. ESI allows rapid, meaningful comparisons across territories that can be tracked as the outbreaks unfold. The ESI quantifies severity relative to a parameterised baseline rather than attempting to estimate values for infection fatality rates, case fatality rates or transmission rates. The relative nature of the ESI sidesteps any problems of confidence associated with absolute rate estimation methods and offers immediate practical strategic value.

## Introduction

COVID-19 has presented strategic decision-making difficulties for countries globally with many territories adopting the same lockdown strategies as countries with severe outbreaks despite there being no evidence of them having similarly severe outbreaks.

Respiratory diseases show strong seasonal patterns [1] varying substantially in summer compared to winter. Transmission rates depends on local weather and environment, and case fatality rates (CFRs) depend on local conditions such as care system quality and capacity, and the general health and immunity of the local population. There is a striking latitude pattern to COVID-19 outbreak severity (see figure 1) with no obvious economic correlation. The data suggest CFRs have varied strongly with latitude, not only transmission rates as might be expected. Given this variance, it is clear that local strategies should be informed by local conditions. A disease with a high CFR may require a ‘suppressive’ strategy (i.e. quarantine, or lockdown), whereas when CFR is low, either naturally or because of available interventions such as vaccines, a ‘mitigation’ strategy is likely to be more effective at reducing total deaths [2] as well as inflicting substantially less economic damage.

**Figure 1.**
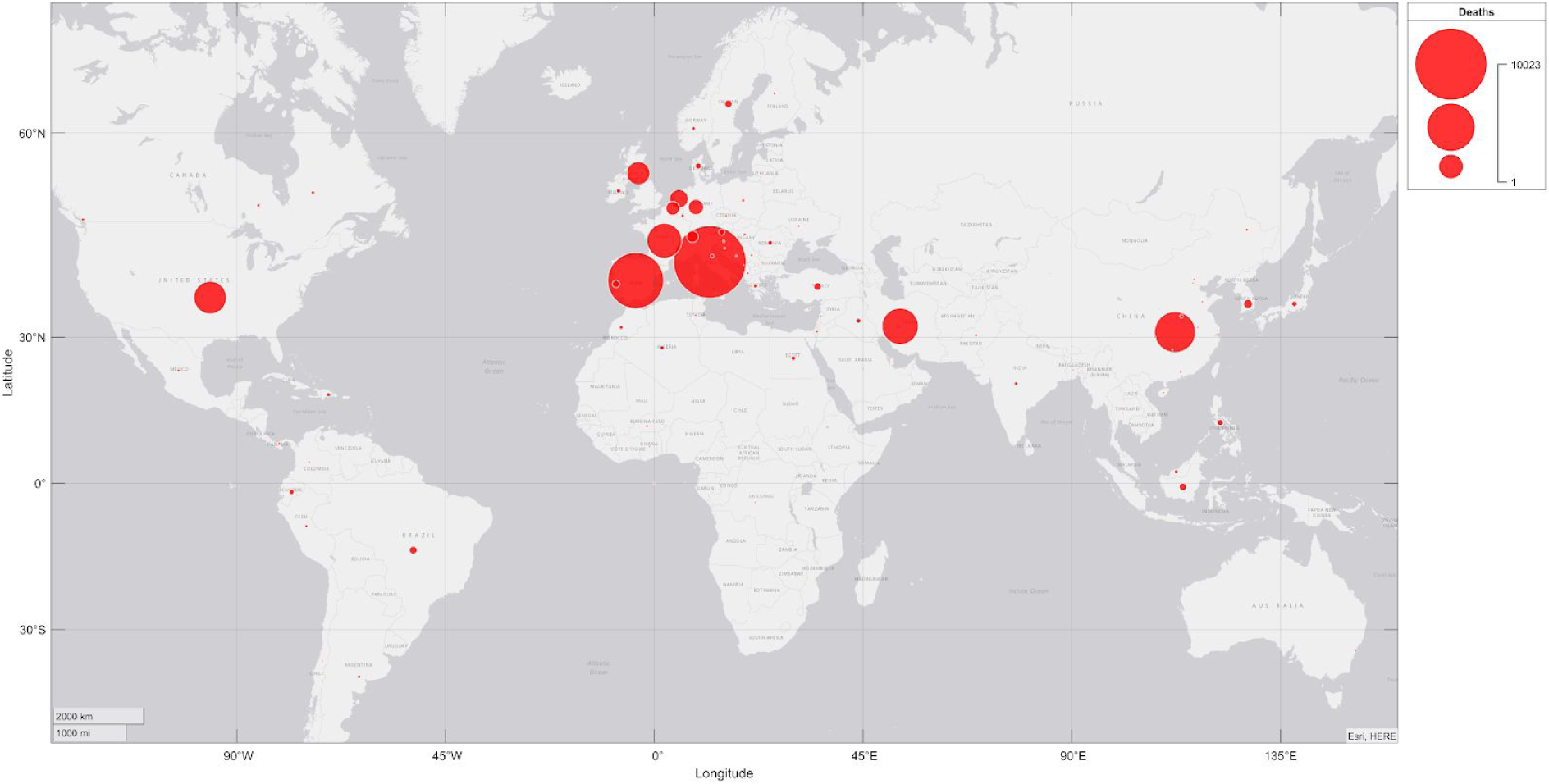
Total COVID19 deaths up to 28 March 2020. Severe outbreaks occurred only in the northern hemisphere.

Looking at deaths alone is not sufficient to determine if an outbreak is severe. Severity assessments must also take into consideration the relative number of recoveries as well as the timing of the outbreak to make a clear judgement of severity.

Various methods exist for estimating novel disease severity, but each has drawbacks that may have hampered their adoption for strategic local decision makers. Many estimates for infection fatality rates and case fatality rates were produced early on in the epidemic but these varied substantially in magnitude [3], with values as low as 0.2% and as high as 28% due to differences in methods and the estimated values for parameter driving them. Many methods define cases from the onset of symptoms but these are then subject to “statistical censoring”, a bias arising from the fact that case figures contain as-yet-uncounted deaths which yet to happen. Methods exist that can adjust for such biases but at the added cost of more complexity and uncertainty. In addition, published global data for COVID-19 did not appear to include cases matching this definition, thus ruling out these methods for quantifying local severity for comparative analysis. Other sources of bias include a shift in case ascertainment over time (due to testing, policy and protocol changes) [4]. Case Fatality Rates (total deaths over total cases) depend on accurate figures for total cases, but in the early days of an epidemic, there is yet another bias towards diagnosis of severe cases leading to severity estimates being overestimated. Even within a single country the case number is a highly volatile metric changing constantly as countries adapt to an exponentially growing problem. It is not possible to compare case counts across countries in any way that is meaningful, but because counts exist people naturally compare them.

The CDC developed their Novel Framework for Assessing Epidemiologic Effects of Influenza Epidemics and Pandemics which is a four-step framework to assess pandemic effects [5]. It uses a complex set of measures (8 for transmissibility and 5 for clinical severity) but its sophistication and reliability on detailed parameters makes it unsuitable for use in rapid global comparisons.

The ability to rapidly quantify and monitor the severity of a new epidemic is clearly of enormous strategic importance: a novel pathogen with a high transmission rate and little or no immune resistance could potentially kill millions within a short time period if a suppressive strategy is not enacted quickly. Conversely, a ‘copycat’ strategy enforcing suppression in regions where mitigation may be the appropriate strategy - simply because other territories did so - not only risks inflicting massive damage to the local economy that is entirely unnecessary, it could potentially lead to worse long-term fatalities as seasons and conditions change: the opportunity to build so-called “herd immunity” when it is most opportune to do so may be missed. As winter in the southern hemisphere progresses, SARS-Cov-2 transmission rates in the south are likely to increase and CFRs are likely to rise. If they reach levels as severe as those seen in the northern hemisphere we may begin to see COVID-19 outbreaks equally severe there appearing from August to September 2020.

### Models and reliability

Traditional transmission models for disease usually depend upon two or more parameters for which ranges of values must be estimated. Models may be made more sophisticated by including new parameters, but whilst each new parameter increases the model’s power to model complex scenarios it simultaneously *decreases* the model’s ability to make accurate forecasts: each new parameter introduces a new source of uncertainty - a new potential source of error. The “parameter space” of a model grows larger in dimension with each parameter and so the requirement for precise and accurate inputs becomes correspondingly greater. A powerful model can ‘fit’ many different disease spread patterns; this is useful for understanding historic outbreaks as well as exploring scenarios, but great flexibility is not a desirable quality for forecasting. Forecasts need to be as clear and precise as possible.

In the early days of a new epidemic simple models with as few parameters as possible are essential. The most useful models depend upon one or two parameters representing the most reliable available evidence. In the first few weeks of an outbreak, tracking deaths over time on a log scale is likely to be the most dependable way of assessing disease severity. A straight line on a log scale indicates exponential growth and it can be quickly established if strong measures are required from this alone.

However, as the epidemic progresses, a reliable method is needed to estimate the severity of outbreaks that uses only reliable early-available data, and that is able to make judgements that take local conditions into consideration.

## Methods

The ESI was developed using data made publicly available on github by John Hopkins CSSE [6]. Some files containing data for deaths and recoveries in CSV format were preprocessed in Excel before being imported into Matlab. Inconsistencies in location definitions between deaths and recoveries data for Canada were reconciled before processing and analysis of the data.

ESI values over time were calculated for territories in six geographic regions using three different baseline severity definitions to show the evolution of the index over time and demonstrate that it is a useful relative measure of severity. Finally, ESI peak values and values at 9th April were plotted as geobubble data using linearised ESI and output logarithmic values and compared with the deaths and recoveries data similarly plotted to demonstrate the utility of this method for comparative analysis.

Data and code are available online at github: https://github.com/gruffdavies/GD-COVID-19.

### Design Principles

The Epidemic Severity Index was designed according to the following principles:

1. **Simplicity** - it generates a single, easily interpretable output value number
2. **Clarity** - it makes a clear distinction between severe outbreaks that require strong measures, and mild outbreaks where different disease management strategies may be more appropriate
3. **Parsimony** - it uses as few parameters as possible as inputs
4. **Reliability** - the evidence parameters representing disease characteristics are chosen based on the likelihood of them being available early, and the most likely to be accurate and precise; no estimated values are required as inputs
5. **Locality** - input parameters and output values reflect local conditions for each country or city
6. **Sensitivity** - it can be used to estimate severity early when fatality numbers are still low
7. **Forecast Capability** - tracked over time the ESI rate of change indicates how likely the outbreak is to become serious and how well-managed the disease is
8. **Adaptability** - ESI has one hyperparameter (an input which is not a local measure of the disease) that allows the specification of the baseline meaning of ‘severe’
9. **Logarithmic** - the range of fatalities may range from tens to millions. Numbers over many orders of magnitude can be hard to compare so ESI output values represent order of magnitude of disease severity, not linear-scale severity.
10. **Robustness** - the index is stable over rapidly changing conditions and is not sensitive to changes that may reflect the changing care and reporting systems rather than the severity of the disease outbreak

### Design Motivation

The severity of COVID-19 outbreaks displayed a striking latitude dependence with severe outbreaks happening almost exclusively above 30°N latitudes, most above 40°N. In severe outbreaks, the reported recoveries and deaths tracked almost 1:1. Below this latitude, outbreaks have been mild and reported recoveries tended quickly to exceed deaths - in some cases by two orders of magnitude. Outliers in the north and south displayed a recoveries-to-deaths pattern matching the general trends of the opposite hemisphere showing that this ratio is a useful determinant of severity. A typical example of this pattern is shown in figure 2.

**Figure 2.**
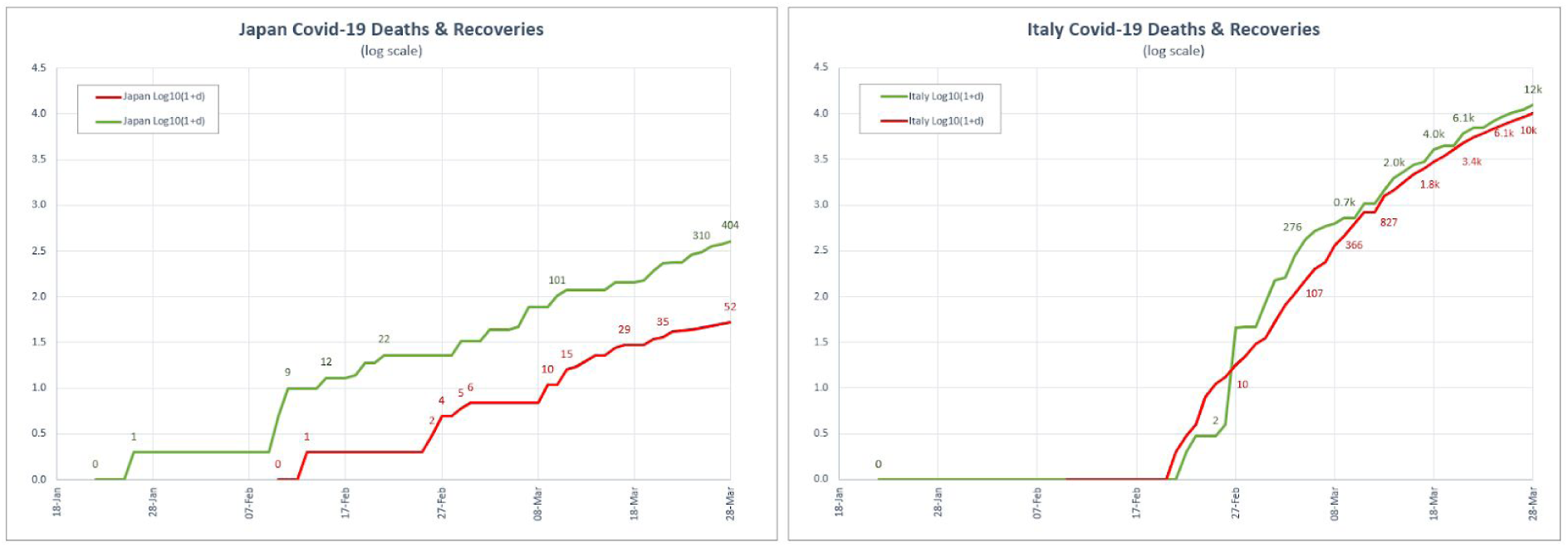
Japan has latitude range comparable to Italy^1^ in the northern hemisphere but its disease progression pattern matched those of economically comparable countries in the southern hemisphere.

### Parameters and Hyperparameters

The Epidemic Severity Index (ESI) uss two parameters and one hyperparameter:

Parameters

*D* Reported number of deaths
*R* Reported number of recoveries

Hyperparameter

*S* Baseline hospital survival ratio (ratio of recoveries to deaths)

The value of *S* defines a local meaning for ‘severity’ to use as the benchmark for judging severity. That is, outcomes that are worse than 1 in S hospital deaths are treated as evidence of “severe” and outcomes that appear better are treated as “not severe”.

### Model Assumptions

ESI assumes
1. Reported deaths come from hospitals and are validated either by tests or clear symptoms in the absence of tests
2. Reported recoveries come from hospitals and represent recoveries from cases severe enough to require hospitalisation
3. Deaths and recovery numbers reflect individuals based in the location (as opposed to visitors whose health is therefore not reflective of local conditions)

As outbreaks become managed and testing capacity increases, assumption 2 is less likely to hold true since recovery reports will no doubt start to come from other sources than hospitals, but this requirement simultaneously becomes less necessary and less important. The utility of ESI is therefore robust to such changes. Nonetheless, interpretation of ESI values should always consider the real contexts determining these inputs.

## The ESI equation

The Epidemic Severity Index, *I*, can be viewed as the product of two terms:

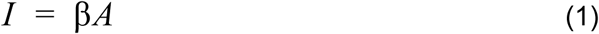

where

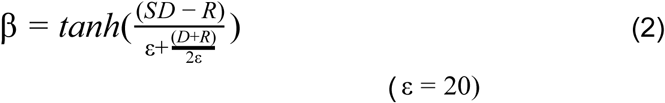

and

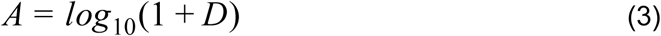

β is a factor that estimates how severe the outbreak appears to be based on the ratio of reported deaths and recoveries using the baseline hyperparameter, *S*, as the definition of ‘severe’. *A* is a measure of the amplitude or scale of the outbreak on the date of the reported values for *D* and *R*.

The hyperbolic tangent function, *tanh*, constrains the severity term, β, to the range -1 to +1. Negative values indicate low severity and positive value indicate high severity relative to the definition of severity supplied by *S*. β is normalised using the total reported hospital cases, and ε is a small ‘stabilising value’ that ensures β is stable for small reported values of deaths and recoveries. A value of 20 was found experimentally to give good results and this can be considered a constant rather than another hyperparameter, though it can be set to any value more than or equal to 1. Higher values improve stability at the cost of reduced early sensitivity, whereas lower values increase sensitivity but decrease stability. ESI will not report useful severity scores until total reported deaths and recoveries exceeds ε.

The scale factor *A* is a logarithmic function of the number of deaths, such that the final output value for ESI is in a low-number range suitable for an index. The logarithmic scale the resulting severity index is intentionally similar to the Richter Scale describing earthquake severity. This is hopefully a familiar concept and it will be understood that 4 is ten times worse than 3, which in turn is ten times worse than 2 etc. In the case of ESI, a value of 1 indicates tens of deaths, 2 indicates hundreds of deaths, 3 indicates thousands, and 4 indicates tens of thousands, and so on.

Unlike the Richter scale, ESI values can intentionally be negative. One of the underlying design principles is that ESI makes a clear distinction between mild outbreaks (given by negative values of ESI) and severe outbreaks that are not currently under control (positive values greater than +1).

The ESI can be plotted using its logarithmic form, or in a linearised form which is useful for geobubble plots where the area of the bubble is proportional to the linearised ESI. Since ESI can be negative, it can be linearised by raising ten to the absolute value (i.e. the ESI stripped of any minus sign). Negative values can be represented by colour to distinguish them from positive values.

### Choosing values for S

Value choices for the hyperparameter, S, were guided by comparisons with published data for seasonal influenza. The US Centers for Disease Control and Prevention reported recent [7] hospitalisation rates of 1.8%, of which 7.5% died (1 in 13), and a final estimate of CFR of 0.14% (see figure 3).

**Figure 3.**
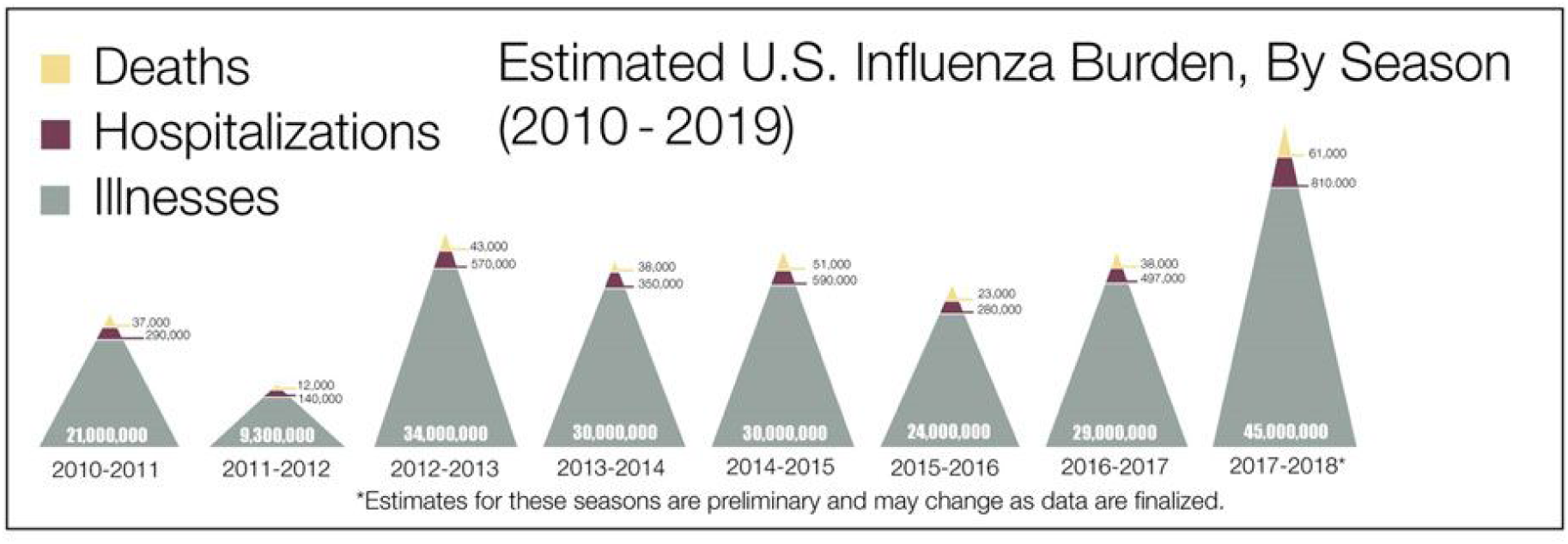
CDC reported illnesses, hospitalisations and deaths for US seasonal influenza

Noting that S scales inversely with severity - it represents recovery not fatality - choosing a value for S *greater* than 13 represents a disease which is *less* severe than seasonal influenza. A choice of *S = 6.5* is equivalent to a disease twice as severe as seasonal flu and *S =* 4.4 corresponds to three times as severe^2^. (See table 1)

**Table 1.** Various choices for S expressed in terms of approximate severity relative to seasonal influenza in the US

| **S** | **Relative severity to seasonal flu** |
| --- | --- |
| 2 | 6.5 times |
| 4.4 | 3 times |
| 6.5 | 2 times |
| 10 | 1.33 times |
| 13 | Equally severe |

In most developed countries that have experienced severe outbreaks the reported recovery and death numbers have tended to track almost 1:1 (i.e. about half of hospital patients survive) until the outbreak was brought under control. This corresponds to S = 2 and suggests COVID19 has a case fatality rate in those countries approximately 6 to 7 times that of seasonal influenza^3^, at around 1%.

Choosing S = 6.5 conditions the ESI formula to judge local outbreaks as severe only if they appear to be approximately more than twice as severe as seasonal influenza.

Informed by the above, ESI was calculated using three different value choices, S = *4.4, S* = *6.5* and *S* = *13.0,*. corresponding approximately to *“at least three times worse”, “at least twice as bad”* and *“at least equally bad”* relative to the severity of seasonal flu in the US respectively.

### Interpretation of ESI values

In the early days of an outbreak the value for ESI may change daily and, assuming the input values can be trusted, a stable rate of change will be more indicative of outbreak severity than the ESI value on any given day.

When the number of reported deaths and recoveries is low the value of ESI is constrained to be in the range -1 to +1 and is likely be volatile^4^, since the ESI is a logarithmic scale this volatility is expected and should not be interpreted as meaningful. ESI values beyond this range they are more stable and meaningful.

### Regions and timing of infections

The start of an outbreak is an important consideration when interpreting the evolution of an outbreak, and ESI time series values were plotted using both absolute dates and days relative to “day zero” herein defined as the first reported death or recovery. This condition was chosen to mitigate any differences in test availability or testing protocols that may have biased first reported infections. In order simplify plots, locations were grouped into six regions defined by geography and the timing of the infection. The timeline of the spread of COVID-19 according to first reported death or recovery was categorised by week (figure 4).

**Figure 4.**
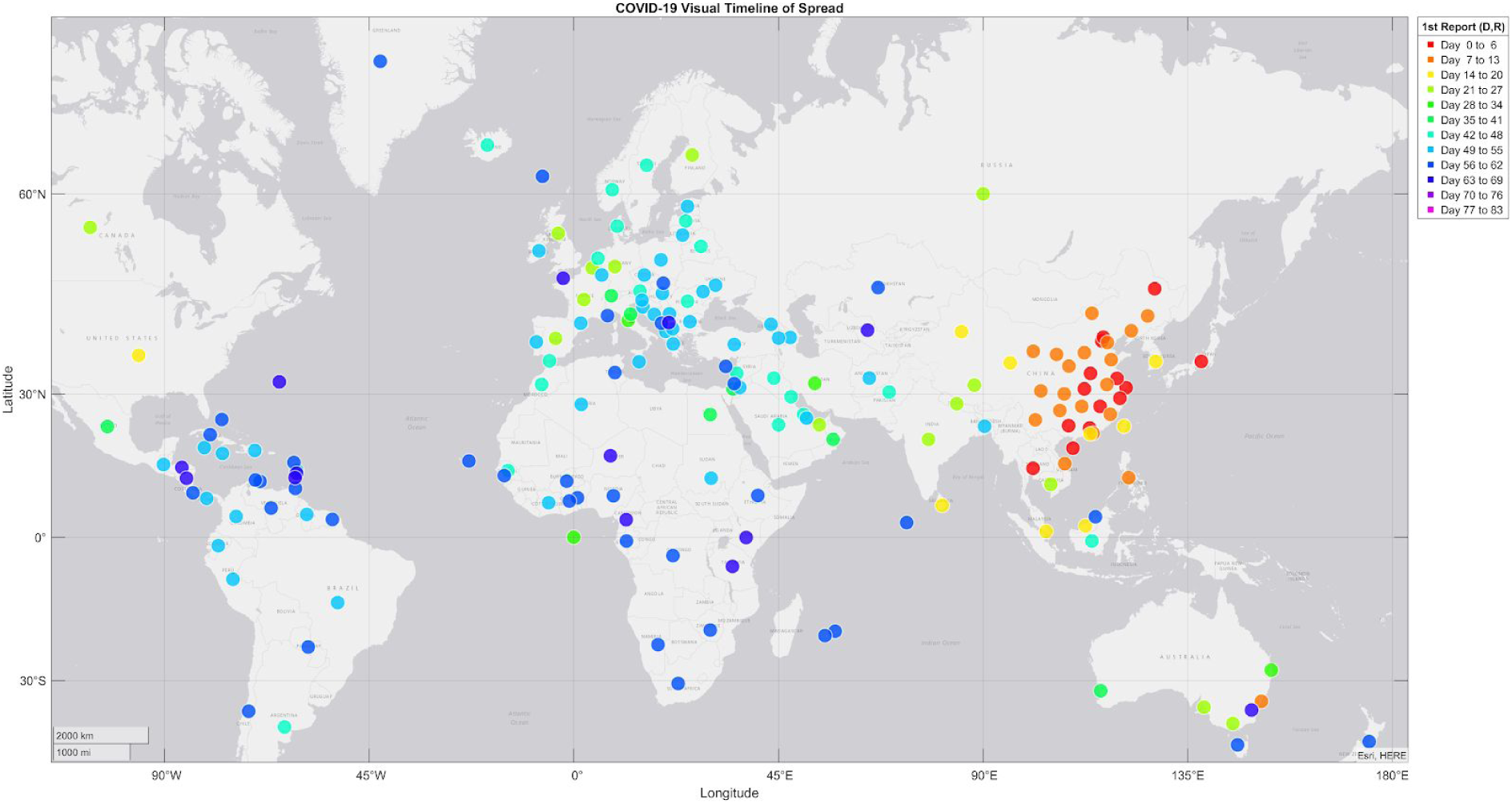
Visual timeline of global spread of COVID-19

This timeline was used to group locations into one of six regions based on geographic region and date of infection. The six regions are:

Region 1: Locations in China where the outbreak started
Region 2: Nearby countries in Asia that were second to report recoveries or deaths
Region 3: Countries in Oceania that were 3rd to report recoveries or deaths
Region 4: Malaysia, Singapore and Sri Lanka (4th to report)
Region 5: Europe, US and Canada
Region 6: South America and South Africa

## Results

ESI time series values were calculated using three different severity parameter values for key locations in the six regions.

Figure 5, figure 6 and figure 7 show ESI values versus date over the range January 22nd to March 28th 2020 for the three choices of *S (4.4, 6.5* & *13.0)*.

**Figure 5.**
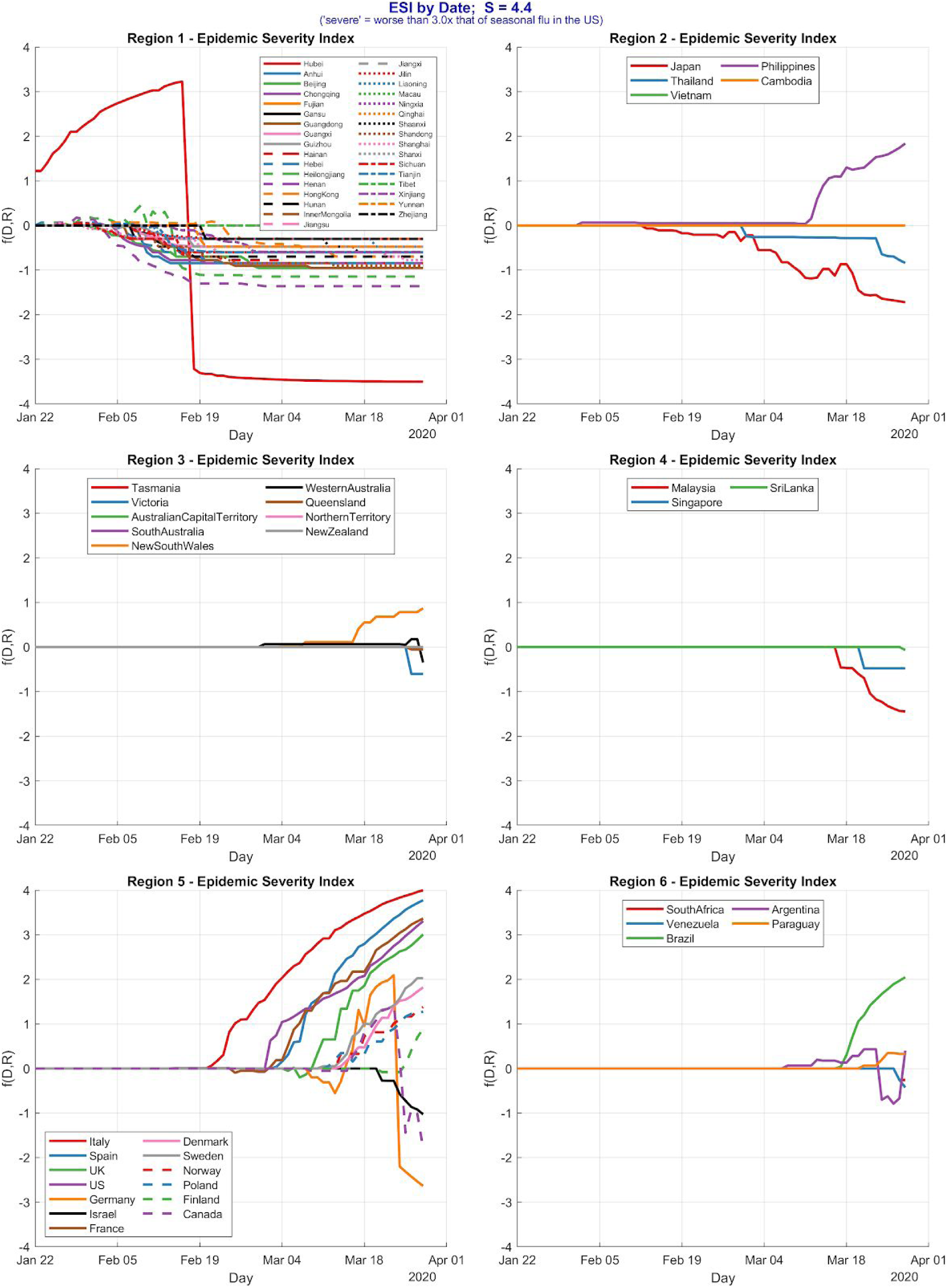
ESI by date(*S* = 4.4) for locations in Six Regions

**Figure 6.**
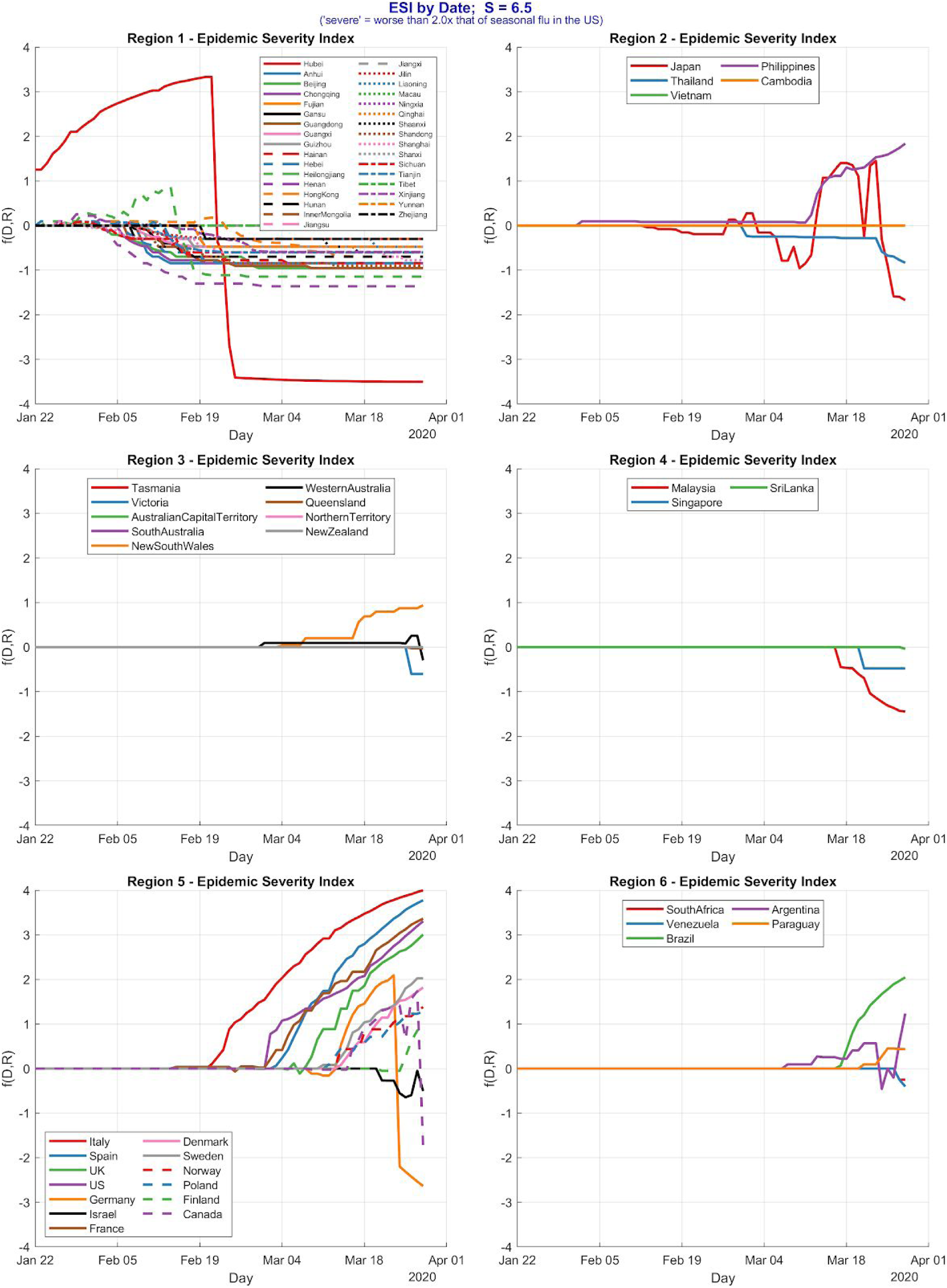
ESI by date(*S* = 6.5) for locations in Six Regions

**Figure 7.**
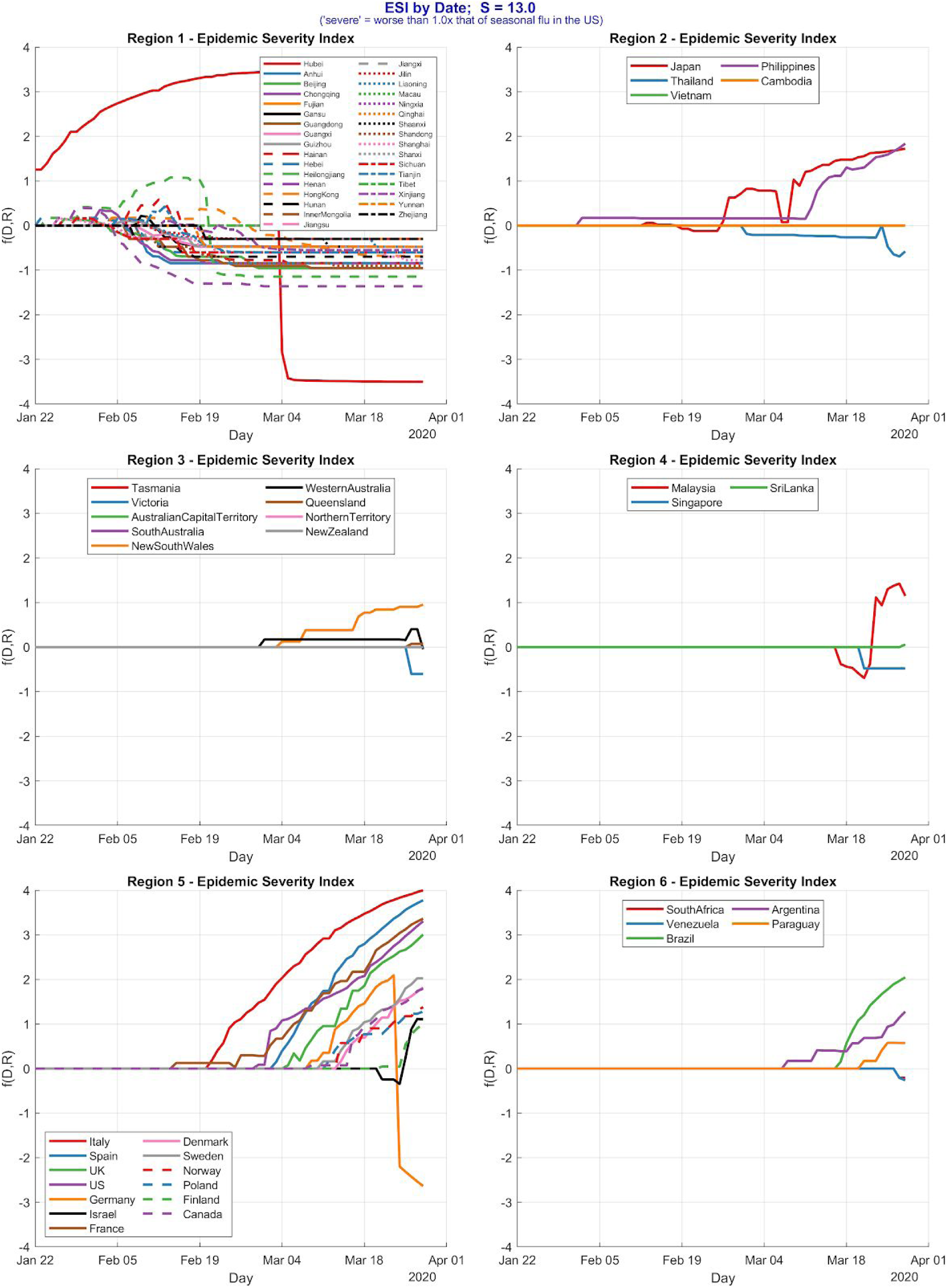
ESI by date(*S* = 13.0) for locations in Six Regions

Figure 8, figure 9 and figure 10 show the same ESI calculations relative to “day zero”. The graphs represent a 70-day range. This relative shift allows for a direct, meaningful comparison of severity across each country and region.

**Figure 8.**
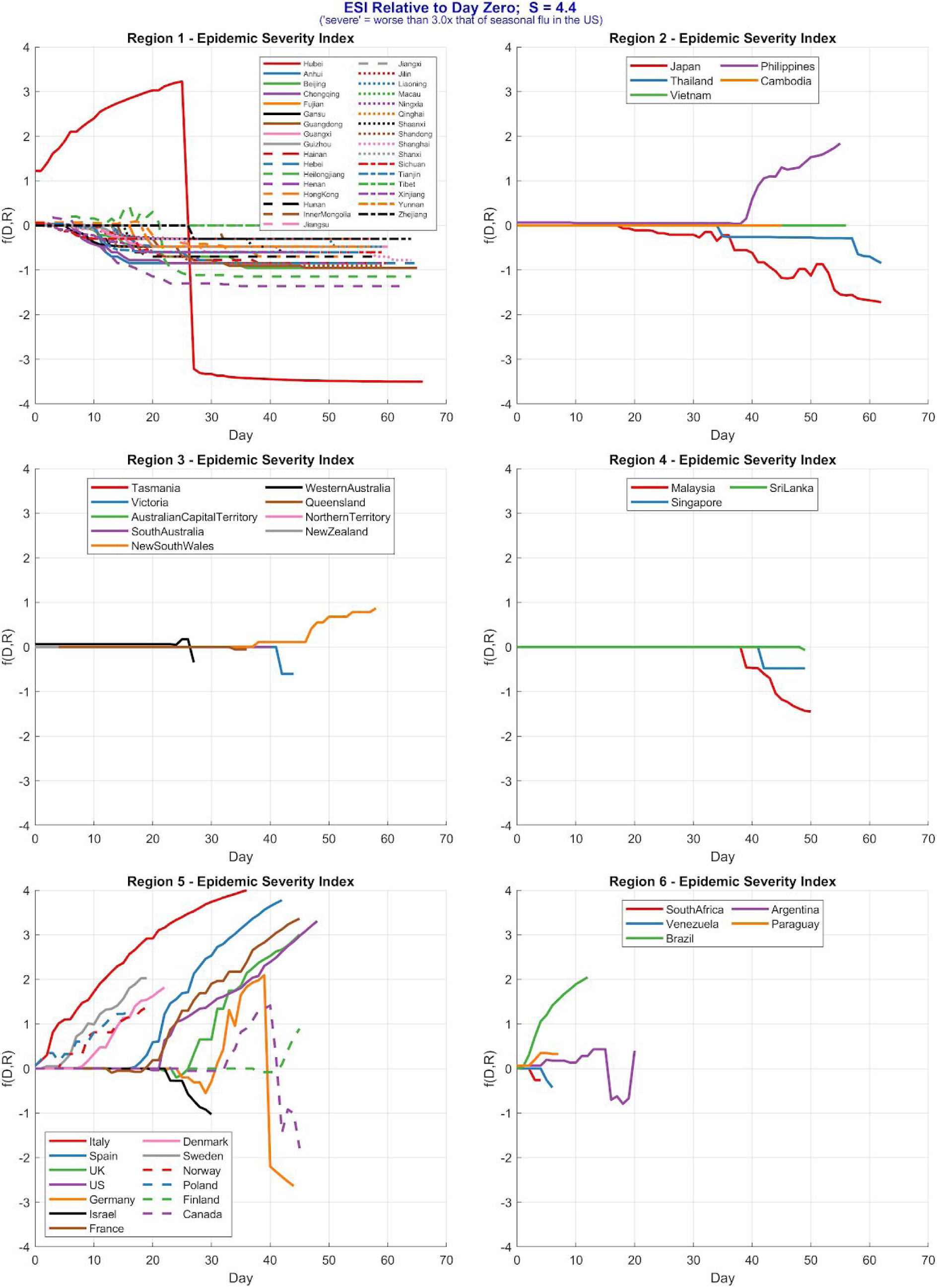
ESI relative to day zero (*S* = 4.4) for locations in Six Regions

**Figure 9.**
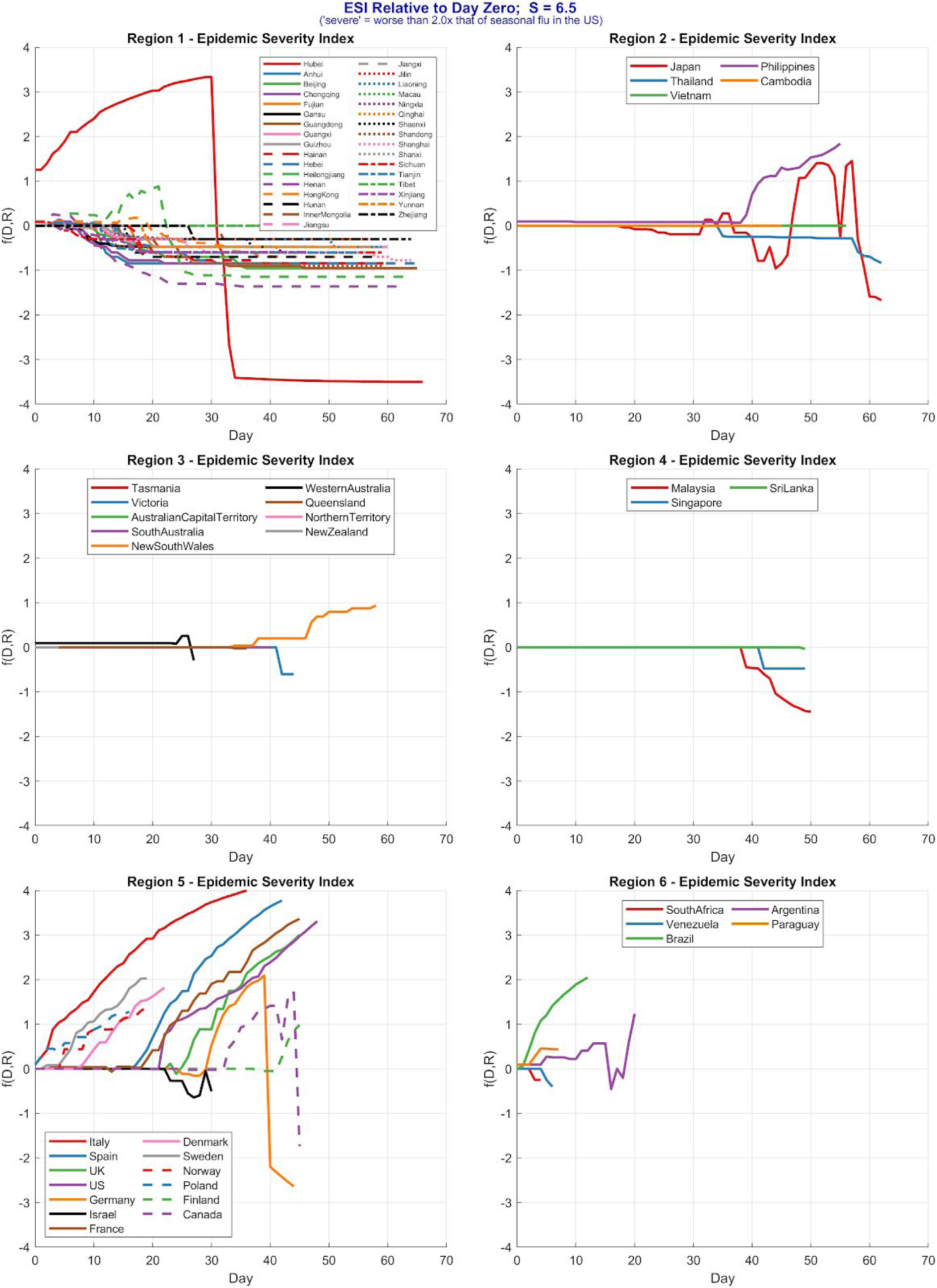
ESI relative to day zero (*S* = 6.5) for locations in Six Regions

**Figure 10.**
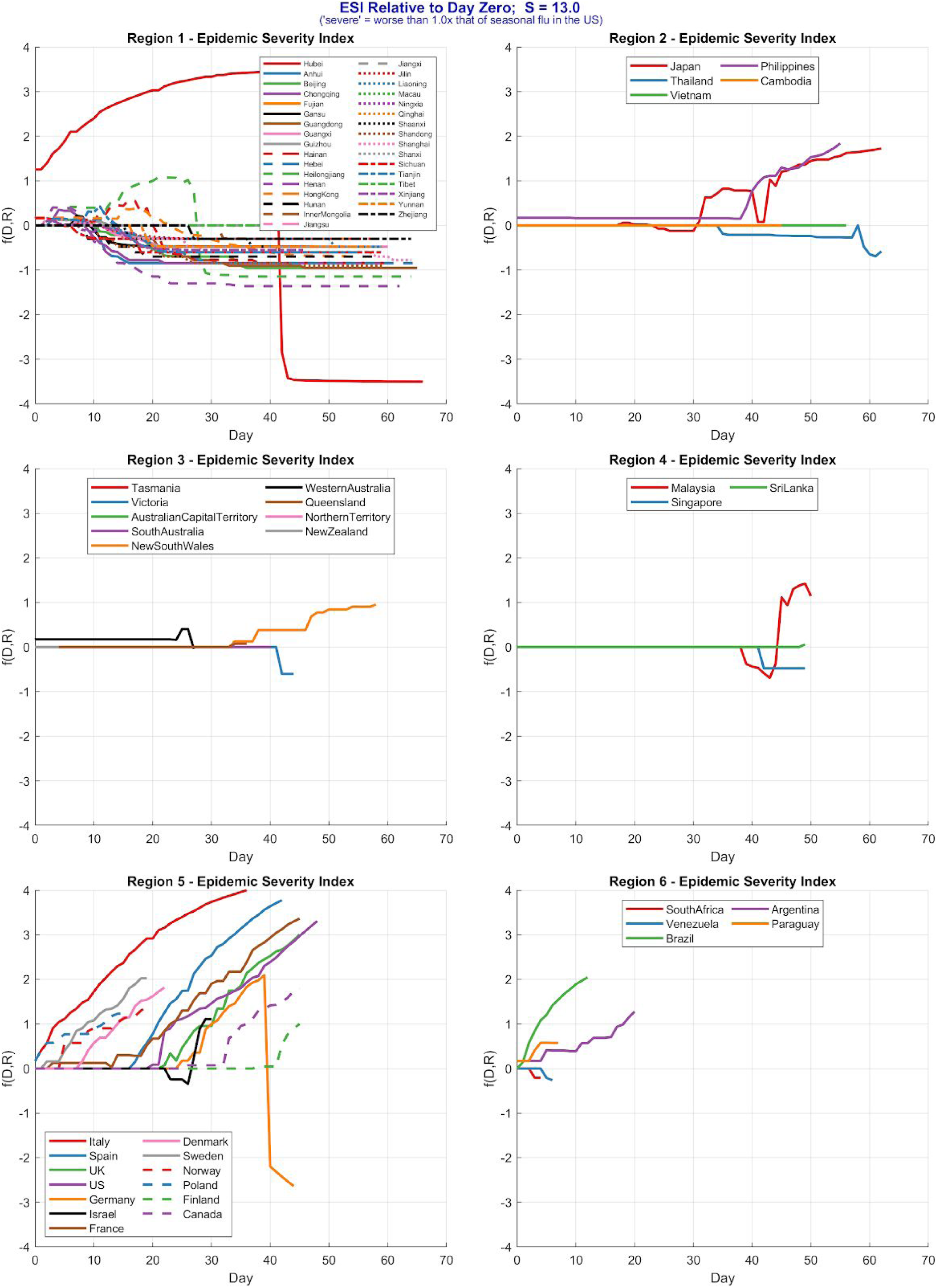
ESI relative to day zero (*S* = 13.0) for locations in Six Regions

### Geobubble plots of ESI, Deaths and Recoveries

The timeseries case studies demonstrated that a useful choice of S = 6.5 for COVID-19. This represents a baseline severity of approximately “twice the severity of seasonal flu in the US”. Total deaths and recoveries are compared here with ESI scores using this value for S.

Since ESI is a logarithmic scale score, it can be useful to express it in a linearised form on a map. This is achieved by raising 10 to the the absolute ESI score (that is, ESI with the sign dropped, if any). Colour is used to distinguish negative, low and positive values.

Figure 11 show death tolls and figure 12 show recoveries using a linear scale where bubble area is proportional to the number of deaths and recoveries on 9th April 2020. Figure 13 shows the same ESI linearised for ease of interpretation and comparison and figure 14 shows unadjusted log output ESI scores for the same date.

**Figure 11.**
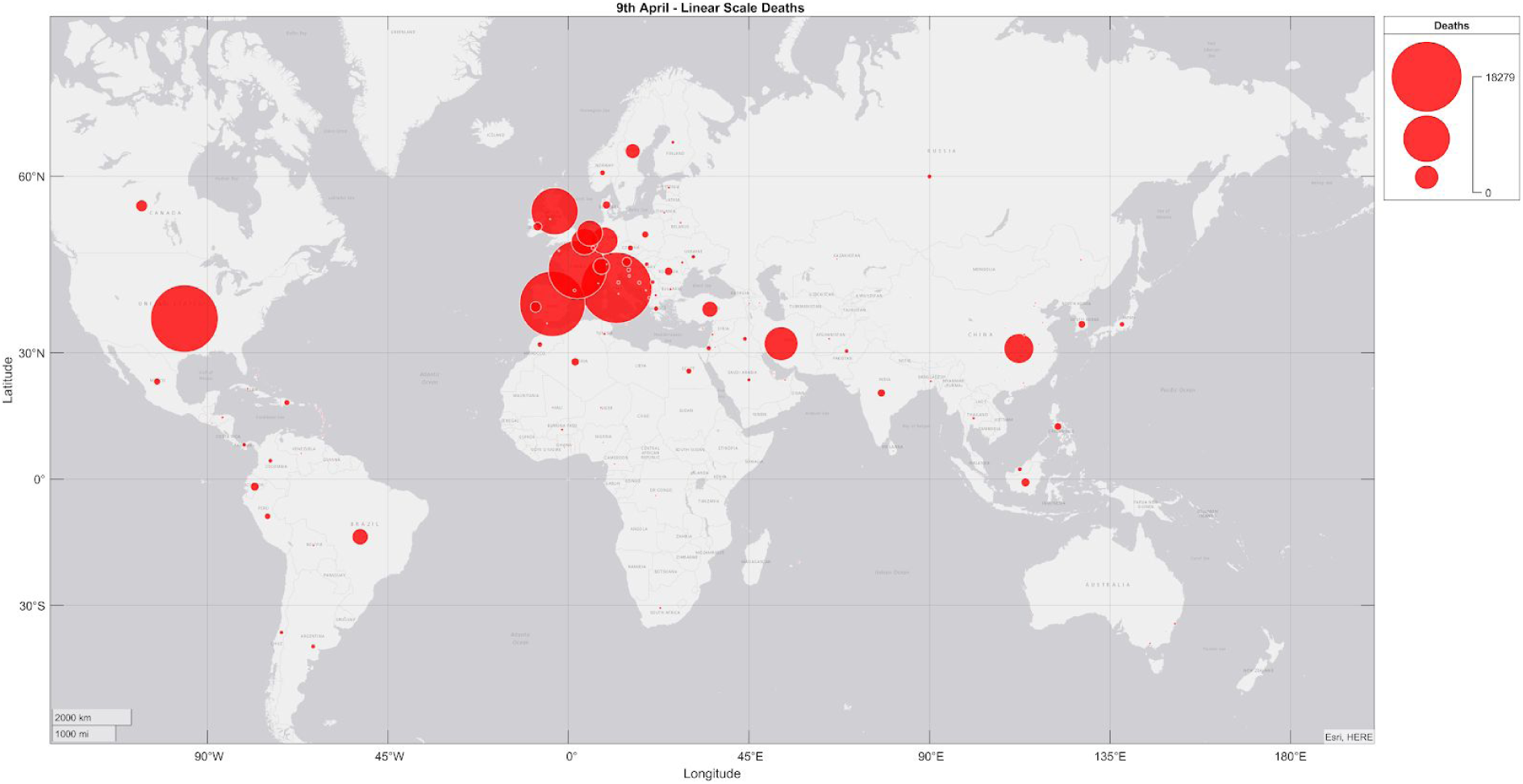
Covid-19 Deaths 9th April (linear scale)

**Figure 12.**
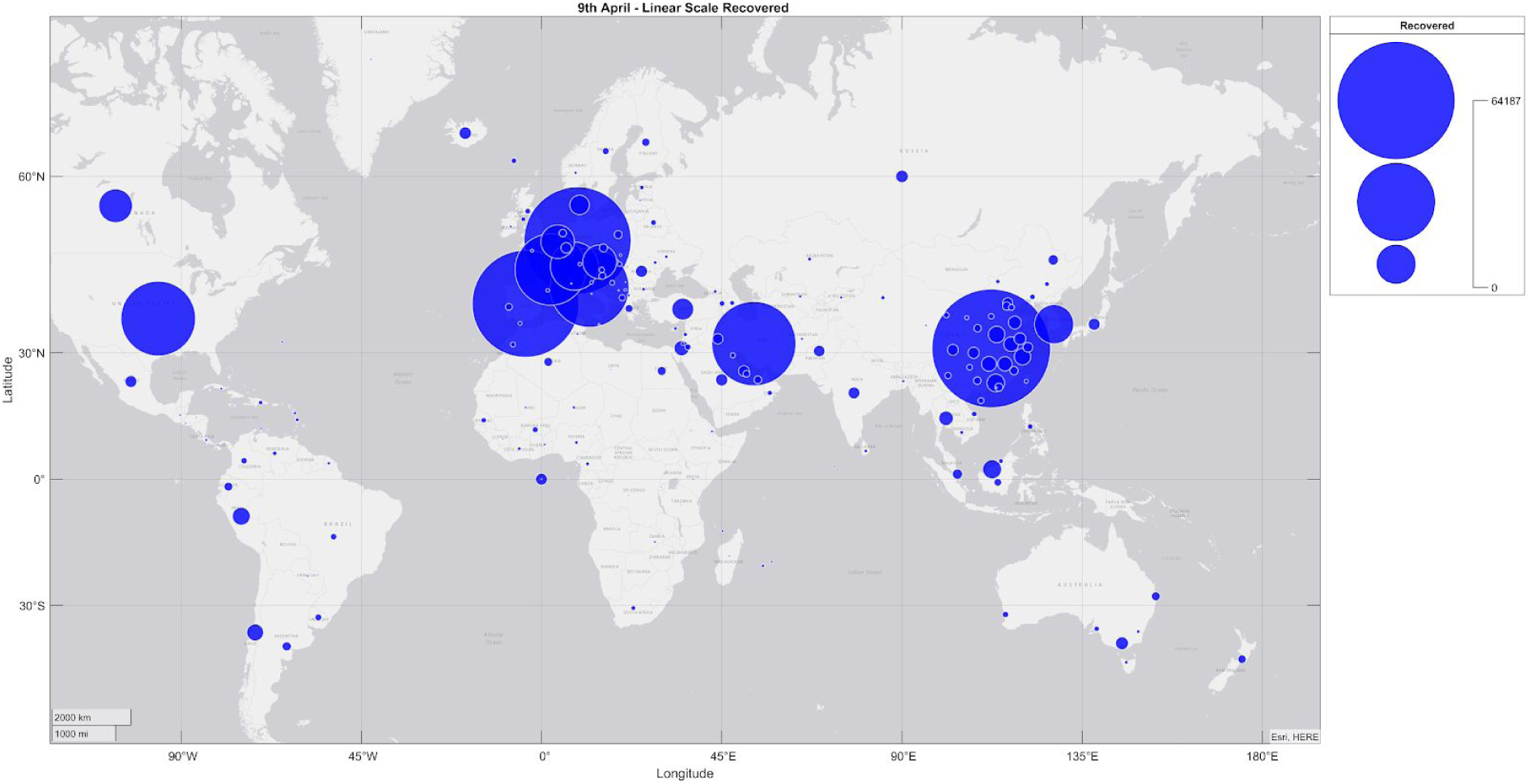
Covid-19 Recoveries 9th April (linear scale)

**Figure 13.**
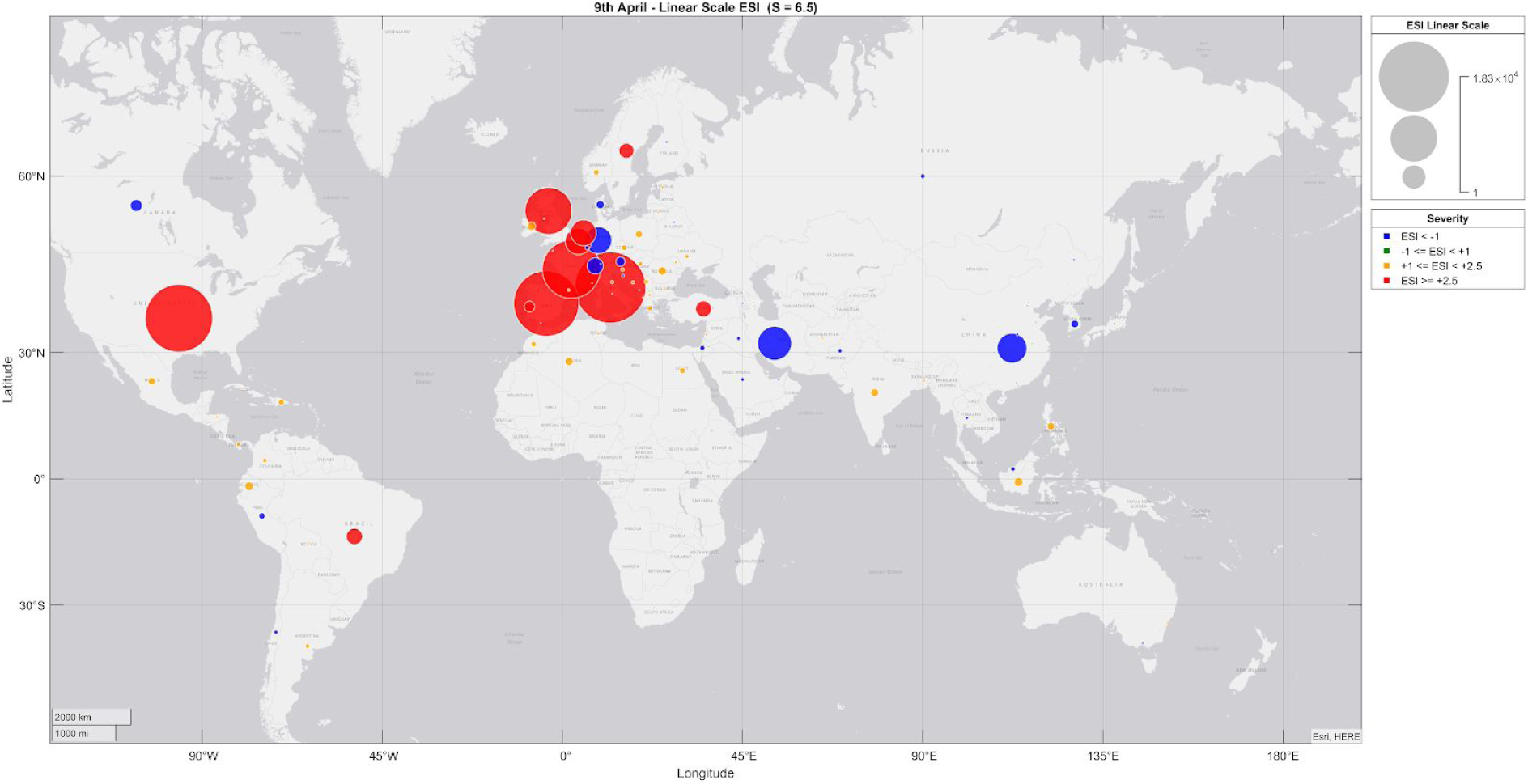
ESI scores linearised for 9th April (S = 6.5) *Colour Key: Blue = ESI < -1; Green = -1 < ESI < +1; Yellow = +1 < ESI < +2.5; Red = ESI > +2.5*

**Figure 14.**
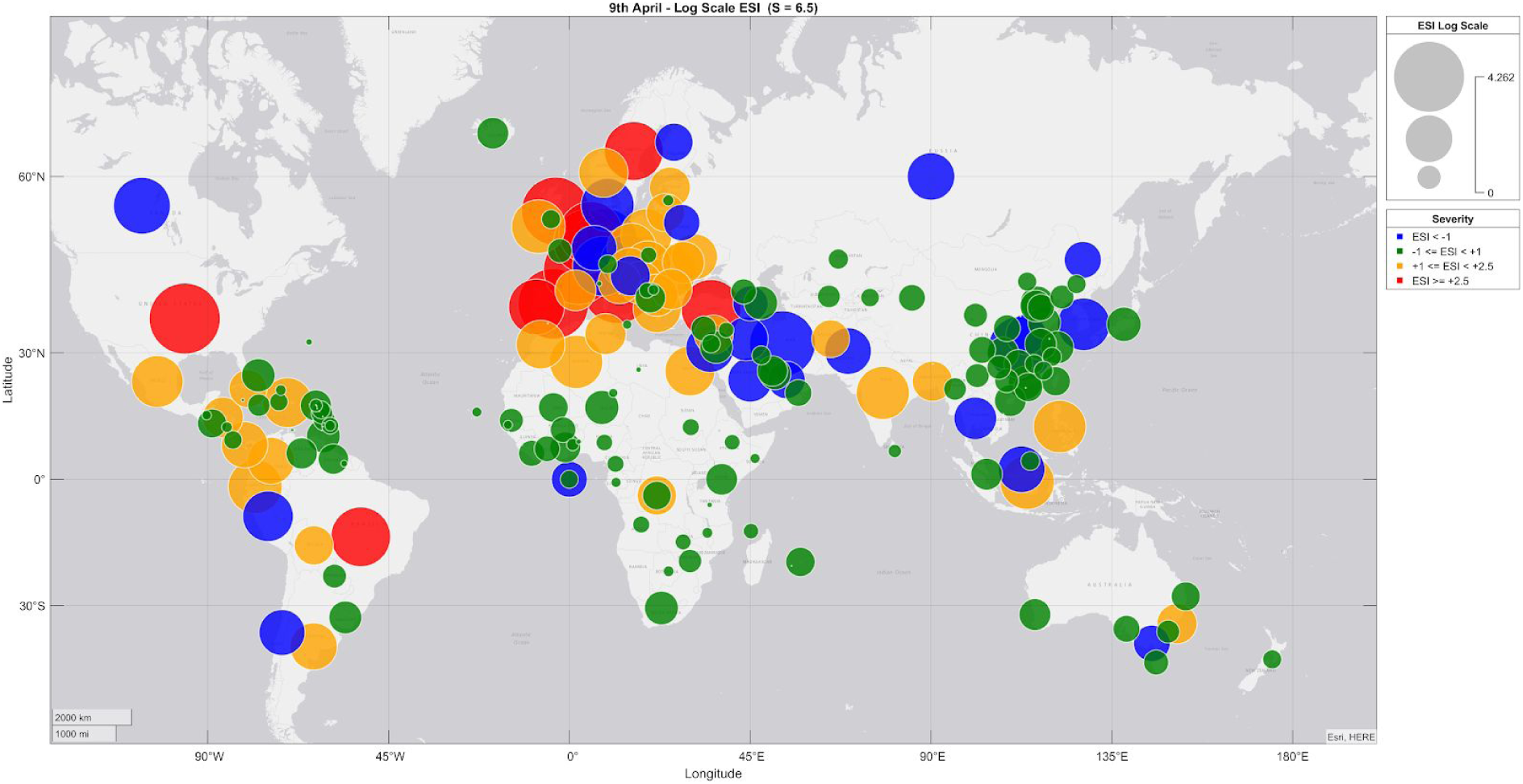
ESI values on 9th April 2020 (S = 6.5; log scale). *Colour Key: Blue = ESI < -1; Green = -1 < ESI < +1; Yellow = +1 < ESI < +2.5; Red = ESI > +2.5*

Finally, figure 15 and figure 16 show the maximum value of ESI attained by March 28th 2020 (linearised and log scale respectively). The cutoff date used was chosen to show the peak severity value of the index up to the end of March marking the start of spring.

**Figure 15.**
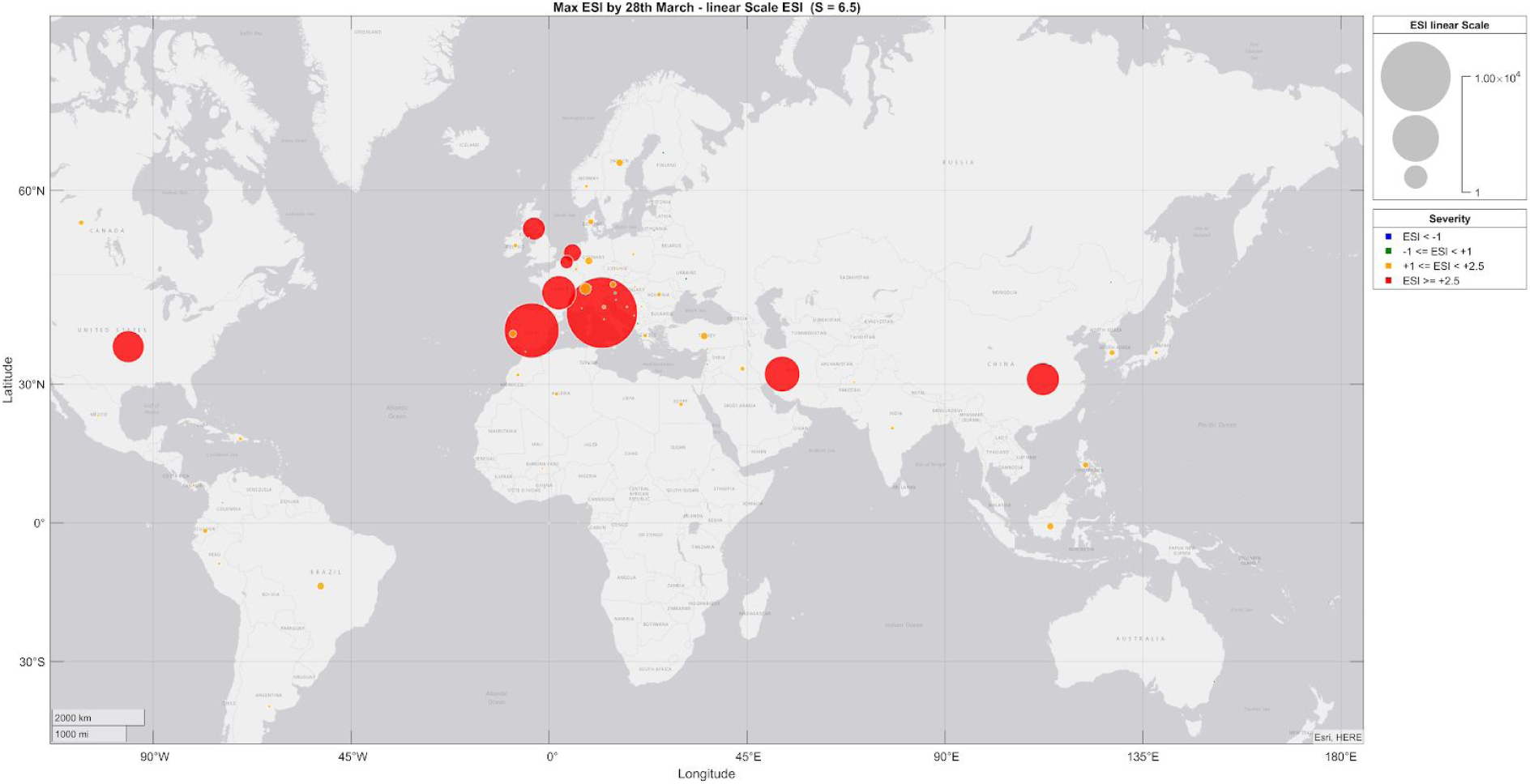
peak ESI value reached up to 28 March 2020 (S = 6.5; linearised scale) Colour Key: Green = ESI < +1; Yellow = +1 < ESI < +2.5; Red = ESI > +2.5

**Figure 16.**
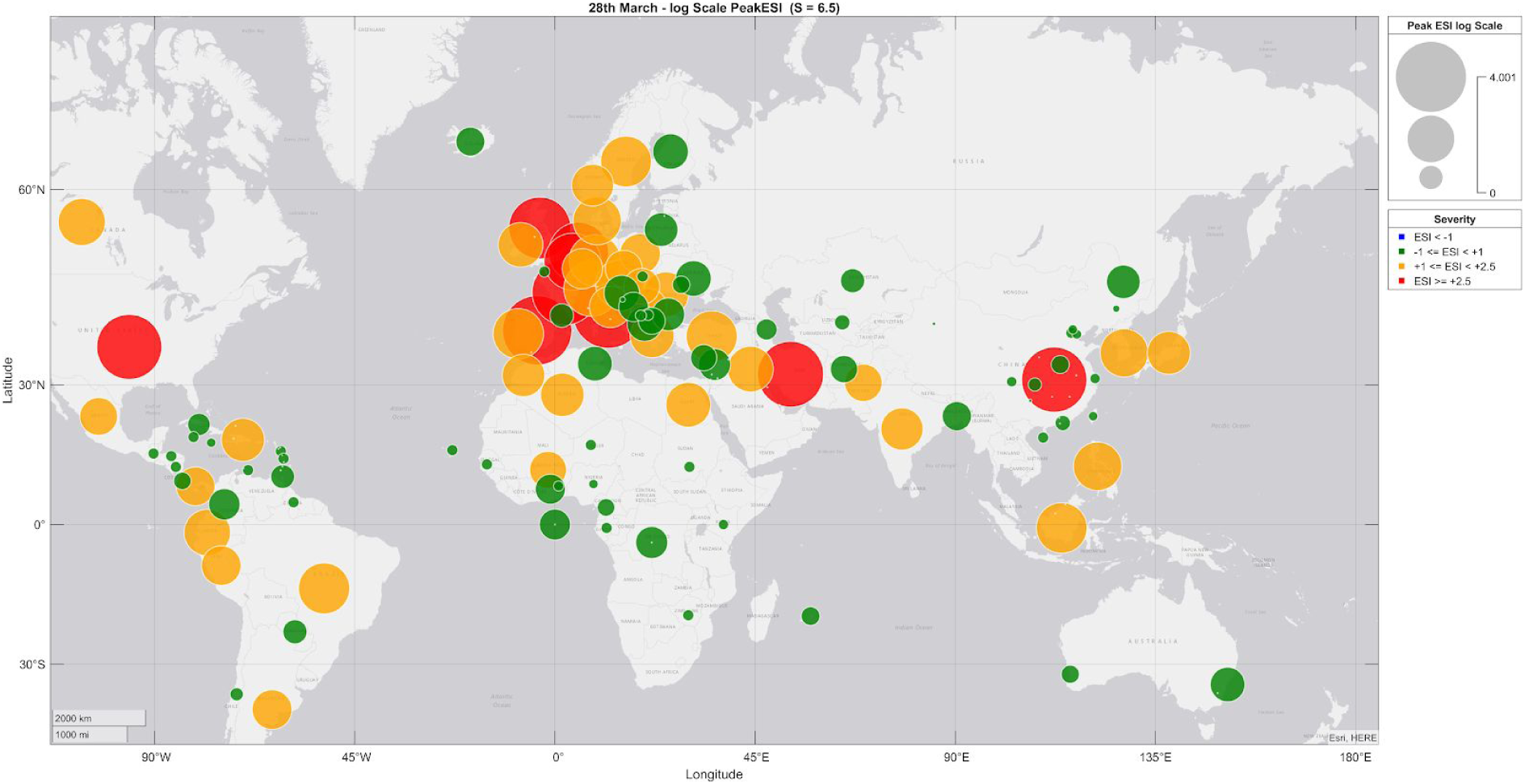
peak ESI value reached up to 28 March 2020 (S = 6.5; log scale) Colour Key: Green = ESI < +1; Yellow = +1 < ESI < +2.5; Red = ESI > +2.5

## Discussion

### Time series plots

The three values chosen for S applied to COVID-19 cases across six regions demonstrate that the large majority of outbreaks have to date occurred in the northern hemisphere. In locations where outbreaks have become severe, infections took hold within weeks of the first reported case. These appear to be confined to locations in Region 1 (China) and Region 5 (Europe and the US).

Territories in Regions 2, 3 and 4 all reported deaths or recoveries before Region 5 and yet severe outbreaks did not develop there in general, with ESI values going immediately negative. In Region 2, the Phillipines appears to be a possible exception to this pattern but up to the 28 March is still only registering an ESI of 2. Brazil appears to be the only country south of the 40°N latitude line where the pattern of outbreak followed a similar trajectory to Regions 1 and 5 in the north.

### Geobubble plots

Plotting ESI using both log and linearised scales is helpful. The default log value scale makes small scores easy to see but these are harder to interpret intuitively. The linearised visualisation re-expresses the index values in a natural scale that offers an more intuitive view. Linearising the scores tends to make scores below +1 vanish as these are very small in comparison to the more severe outbreaks but results in a very clear view of severe-as-yet-uncontrolled outbreaks (red) and severe-but-now-controlled outbreaks (blue).

The log scale plot (figure 14) shows the current situation for each territory with great clarity, but bubble sizes may be misinterpreted on a log scale, so the linearised plot (figure 13) makes a useful reference. Similarly, the peak value ESI plots (figure 15 and figure 16) show the overall severity of outbreaks up to the end of winter in the northern hemisphere. The linearised plots in particular (figure 13 and figure 15) highlight the striking latitude dependence of COVID-19 outbreak severities.

The small number of outliers that break this latitude relationship remain to be explained. Whilst a full investigation of these is beyond the scope of this document, a brief discussion is merited.

### Northern Outliers

Japan is the most striking outlier in the north. It was one of the earliest locations to report detected infections and recoveries yet has experienced very few deaths in absolute terms but also a very low case fatality rate with recoveries as shown earlier in figure 2. Japan has a very high population density with a population of 126 million people and also a large subpopulation of elderly. A third of Japanese population are aged 60 or older [8]. The population density in Japan is 347 people per Km^2^, the total land area is 364,555 Km^2^ and 91.8 % of the population is urban. The median age in Japan is 48.4 years. Excluding small islands, Japan’s latitude ranges from 30°N to 47°N. This compares with Italy which has a latitude range from 37°N to 47°N, has a population of 60.5 million living in an area of 294,140 Km^2^, giving a population density in Italy of 206 people per Km^2^. 69.5 % of the Italian population is urban and the median age in Italy is 47.3 years [9].

Germany has reported data that suggests an outbreak far less severe than neighbouring European countries Italy, Spain, UK and France, and further away, the US, which have all reported severe outbreaks. Germany appears to have brought its outbreak under control quickly. The shape of the evolution of the ESI for German and Japan are interesting to compare in the case where *S* = *6.5*. Looking ESI plotted relative to day zero, (figure 9), both Japan and German display late onset with Germany’s outbreak taking holding around day 30 and Japan’s after day 45, however prior to this, the ESI values first go negative, indicating that early recoveries were reported long before significant numbers of deaths. In both cases, outbreaks then briefly took hold with ESI turning positive, but both countries managed to control the outbreaks over the next 10 to 15 days, with ESI values rapidly going negative again.

Israel’s outbreak appears as mild from the beginning for all three values of *S* but its latitude 31°N places it on the border of the north south divide though this is notably similar to that of Hubei and Iran (latitude 32°N) both of which suffered severe outbreaks.

### Southern Outliers

Two territories below 40°N latitude appear to be cause for concern: Brazil and the Philippines, but compared to the ESI curves for outbreaks in the north, they are much less severe by comparison and the ESI values taper off more quickly. All other locations in the southern hemisphere are consistent with outbreaks that are currently about as severe as seasonal flu.

### Caveat Interpretor

The somewhat paradoxical goal of a high-level statistic is to distil a complex-hard-to-understand picture into a simple-easy-to-understand one whilst faithfully retaining salient information. As is true with any picture, reducing complexity (“resolution”) always comes at the direct cost of losing detail. Any picture reduced to a single point has necessarily lost most of the detailed information. It is therefore vitally important to check that the remaining datum usefully reflects that which has been discarded in the summarisation process. Whilst the ESI has been designed with stringent core design principles, the numerical values that it generates must still be interpreted with great care. As with all formulae, the quality the output depends on the quality of the inputs, and errors or differences in reported deaths and recoveries may account for differences in ESI.

## Conclusions

The ESI provides a simple, evidence-based method for quantifying the relative severity of epidemic outbreaks which uses reported data most likely to be available early for severe cases requiring hospitalisation. These, unlike infection cases, which depend on test availability and protocols, are likely to be consistent and comparable across territories. ESI is able to make meaningful comparisons of severity globally. It does not suffer typical statistical censoring bias. Delay bias that may occur from recoveries taking longer than deaths relative to the onset of infection will bias the severity index higher, which is consistent with longer time to recovery and is therefore desirable.

The ESI reduces the potential for confusion and uncertainty inherent in methods that estimate values for transmission rates and case fatality rates and the large range of values these estimates inevitably produce. The relative nature of the Index allows immediate concrete comparisons across territories. The parameterised nature of the baseline definition for severity provides flexibility and, even though CFR estimates are not involved, this makes it possible to make approximate comparisons with diseases with known CFRs.

The ESI is adds to the available suite of severity estimation techniques offering a level of clarity and ease-of-interpretation that will hopefully make it of immediate and high strategic value to decision makers.

## Data Availability

Data and code are available online at github.

https://github.com/gruffdavies/GD-COVID-19

## Acknowledgements

I’m grateful to the following people for their helpful comments and early feedback: Dr Joanne Byers, Dr Attila R Garami, and Professor Andrea Giustina.

## About the author

Dr Gareth Davies has a BSc in Physics and a PhD in Medical Physics from Imperial College, London, though he is not currently affiliated with Imperial College as a research scientist. He has more than three decades of experience of complex data analysis, systems modelling, software engineering and machine learning. In 2019, he was named as one of Codex World’s Top 50 Innovators.

## Footnotes

1 Excluding small islands, Japan ranges from 30°N to 45.5°N and Italy from 35°N to 47.5°N.

2 If this inverse relationship is confusing, S can be calculated alternatively using 13/F, where F denotes hospital fatality relative to seasonal flu as shown in the right column of Table 1.

3 The fact that a severity of 6.5 times that of seasonal flu is enough to generate greater than 10% hospitalisation rate and 50% hospital survival rate is a sobering reminder of the severity of influenza.

4 A useful metaphor is camera zoom: the ESI equation applies an automatic zoom (scaling factor) in order to ‘see’ smaller numbers in early stages and any ‘shakes’ in the numbers are magnified. As reported numbers grow, the equation ‘zooms out’ and these shakes or instabilities diminish.

